# Purchasing high-cost medical equipment in hospitals: A systematic review

**DOI:** 10.1101/2021.11.10.21266152

**Authors:** Saba Hinrichs-Krapels, Bor Ditewig, Harriet Boulding, Anastasia Chalkidou, Jamie Erskine, Farhad Shokraneh

**Affiliations:** Faculty of Technology, Policy and Management, Delft University of Technology, Delft, The Netherlands; The Policy Institute, King’s College London, London, UK; King’s Technology Evaluation Centre (KiTEC), London Institute of Healthcare Engineering, School of Biomedical Engineering and Imaging Sciences, Faculty of Life Sciences and Medicine, King’s College London, London, UK

**Keywords:** Purchasing, procurement, high-cost equipment, medical devices, hospitals, systematic review, materials management

## Abstract

**Objectives:** To systematically review academic literature for empirical studies on any processes, procedures, methods or approaches to purchasing high-cost medical equipment within hospitals in high-income countries.

**Design:** Systematic review

**Methods:** On 13 August 2020, we searched the following from inception: Cost-Effectiveness Analysis Registry, EconLit and ProQuest Dissertations & Theses A&I via ProQuest, Embase, MEDLINE, and MEDLINE in Process via Ovid SP, Google and Google Scholar, Health Management and Policy Database via Ovid SP, IEEE Xplore Digital Library, International HTA Database, NHS EED via CRD Web, Science Citation Index-Expanded, Conference Proceedings Citation Index-Science, and Emerging Sources Citation Index via Web of Science, Scopus, and Zetoc conference search. Studies were included if they described the approach to purchasing (also known as procurement or acquisition) of high-cost medical devices and/or equipment conducting within hospitals in high-income countries between 2000-2020. Studies were screened, data extracted, and summarised.

**Results:** Of 9437 records, 24 were included, based in 12 different countries and covering equipment types ranging from surgical robots to MRI scanners and orthopaedic implants. Study types included descriptions of processes taking place within or across hospitals (n=14), out of which three reported cost savings; empirical studies in which hospital records or participant data were analysed (n=8), and evaluations or pilots of proposed purchasing processes (n=2). Studies mainly highlight the importance of multidisciplinary involvement (especially clinical engineers and clinicians) in purchasing decision-making to balance technical, financial, safety and clinical aspects of device selection, and the potential of increasing evidence-based decisions using approaches ranging from hospital-based health technology assessments, ergonomics, to conducting user ‘trials’ of the device in use before purchase.

**Conclusions:** We highlight the lack of rigorous empirical work on this topic, calling for more intervention based and empirical work to advance the evidence base in this domain to advance knowledge, policy and practice.

**Strengths and limitations of this study:**

- First systematic review of empirical work conducted in hospitals on purchasing of high-cost medical devices
- Broad search covering a range of disciplines and study types
- Limited to high-cost equipment which is challenging to differentiate across studies and has no standardised ‘value’ globally

## INTRODUCTION

### Context

According to the World Health Organisation (WHO), medical devices and equipment are essential for maintaining health system performance.[1] Inadequate selection and distribution of technologies can create inefficiencies and waste,[2] or create risks to quality of health services, such as in a pandemic.[3,4] To avoid these risks, a large body of literature concentrates on designing devices for patient safety, while other studies have focussed on adhering to regulatory requirements to ensure devices are safe enough for the market. Following this, devices may be evaluated to understand its impacts in specific healthcare contexts and compared against available alternatives, which encompass the field of Health Technology Assessment (HTA).[5] However, there has been less attention paid to the next steps: acquiring, purchasing or procurement of these devices by the health system.

Medical device purchasing, more comprehensively known as procurement, goes beyond basic contracting between the supplier and health provider; it requires consideration of user needs, technical maintenance, training needs, adequate consumables, and how they can be disposed.[6] Despite the potential role purchasing processes play in promoting patient safety[7,8] and efficiency,[9] studies suggest these are not optimised for efficiency and quality. A study comparing medical device purchasing across five countries found that there is more focus on cost-containment, and less on quality and health outcomes.[10] Empirical studies of purchasers in UK hospitals have shown that there are a wide range of stakeholders potentially involved in purchasing decisions (from clinicians, nurses, biomedical engineers, finance staff and/or managers), but their responsibilities and protocols are ill-defined, their skills and expertise differ,[11] they often work in silos and make decisions under high pressure conditions,[12] and that the lack of stakeholder analysis as part of purchasing planning processes resulted in conflicts and delays in decisions.[13] A more recent scoping literature review of the logistics function in hospitals demonstrated that logistics functions can be highly inefficient and fragmented.[14]

### Need for this review

Understanding purchasing processes can help us uncover why some of these inefficiencies and tensions exist, by exploring the inner workings of the environment, protocols, behaviours and organization of purchasing staff and departments, and thereby identifying areas for improved practices. In this review, we sought to identify studies that specifically focus on the purchasing of high-cost medical equipment in hospitals, in high-income settings. Specifically, this meant identifying any process, procedure, method, or approach used within a hospital to reach decisions about which equipment would be purchased. While there are reviews of good practice in purchasing and supply chain management and their applications in health care settings generally,[15,16] to our knowledge there are no comprehensive reviews that demonstrate existing approaches, practices and methods used for purchasing of medical devices and equipment in hospitals specifically in high-income settings. The most similar existing reviews that we found so far include a review of methods for procurement of medical devices and equipment focussing exclusively on low- and middle-income countries,[17] a realist review of theoretical and empirical literature on procurement and supply chain management practices more generally,[15] and a rapid evidence assessment of literature with lessons from the non-health sector to inform health purchasing and supply chain management.[16] None of these systematically searched for academic studies that focussed on the internal workings of a hospital to identify current practices and understand purchasing behaviours, processes and approaches. Two exceptions which do cover activities within hospitals, but with a different scope, are the review by Volland et al 2017[18] which examined studies covering materials management and logistics in hospitals, but with a focus on quantitative methods, and Trindade et al 2019 who focussed on the qualitative assessment of devices, not the process of procurement as a whole.[19]

### Objective and scope of the review

Our research question in this review is framed as: What does the academic literature tell us about the way in which high-cost equipment is purchased in hospitals in higher income settings?

Our review focuses on the steps in hospitals that occur after any HTA exercise, whether it was national- or hospital-based. Medical device purchasing sits within other activities in hospitals, including: health technology management, materials management, supply chain and logistics. Our focus is on what is commonly termed the acquisition process, which begins the moment the need for a new or replacement device is identified, to the moment it is installed and ready for operation. For a comprehensive view of how the medical device and equipment purchasing function of a hospital fits within its wider activities, we refer readers to the WHO procurement process guide.[20]

## METHOD

We followed Cochrane Collaboration’s methods in conducting this systematic review [21] and complied with Preferred Reporting Items for Systematic Reviews and Meta-Analyses (PRISMA).[22] The full protocol for this systematic review is published elsewhere[23] and summarised below.

### Search methods

On 13 August 2020, we searched the following from inception: Cost-Effectiveness Analysis Registry, EconLit and ProQuest Dissertations & Theses A&I via ProQuest, Embase, MEDLINE, and MEDLINE in Process via Ovid SP, Google and Google Scholar, Health Management and Policy Database via Ovid SP, IEEE Xplore Digital Library, International HTA Database, NHS EED via CRD Web, Science Citation Index-Expanded, Conference Proceedings Citation Index-Science, and Emerging Sources Citation Index via Web of Science, Scopus, and Zetoc conference search. An information scientist designed, tested, revised, and ran the searches in collaboration with the review team. The search consisted of three main blocks of setting, product, and process. All search strategies for all sources are reported in Appendix 1.

### Eligibility criteria

#### We included the studies if they met the following criteria

***Process* :** The study describes the process for the purchase (also known as procurement or acquisition) of high-cost medical devices and/or equipment; ***Setting:*** The study setting is one or more hospitals or departments within the hospital(s) in high-income countries (using OECD countries as a proxy indicator for high-income); ***Product:*** The purchased product is a single or a group of high-cost (also known as high-value or capital) medical devices or equipment; ***Practice:*** Studies conducted between 2000-2020 to represent ‘current’ processes reported in hospitals. Studies not demonstrating influence on purchasing decisions or theoretical models not assessed, piloted or evaluated in hospital settings were excluded.

### Study selection

We used EndNote to remove the duplicates and Rayyan for screening the titles and abstracts. Two independent reviewers piloted the screening based on eligibility criteria before conducting a sensitive screening. Two independent reviewers re-screened these relevant/possibility relevant records from sensitive screening and resolved the disagreements in weekly group meetings. We followed dual-screening and arbitration by a third reviewer for the full text screening step. We recorded and reported the reasons for exclusion for any excluded paper at full text stage (Figure 1).

**Figure 1.**
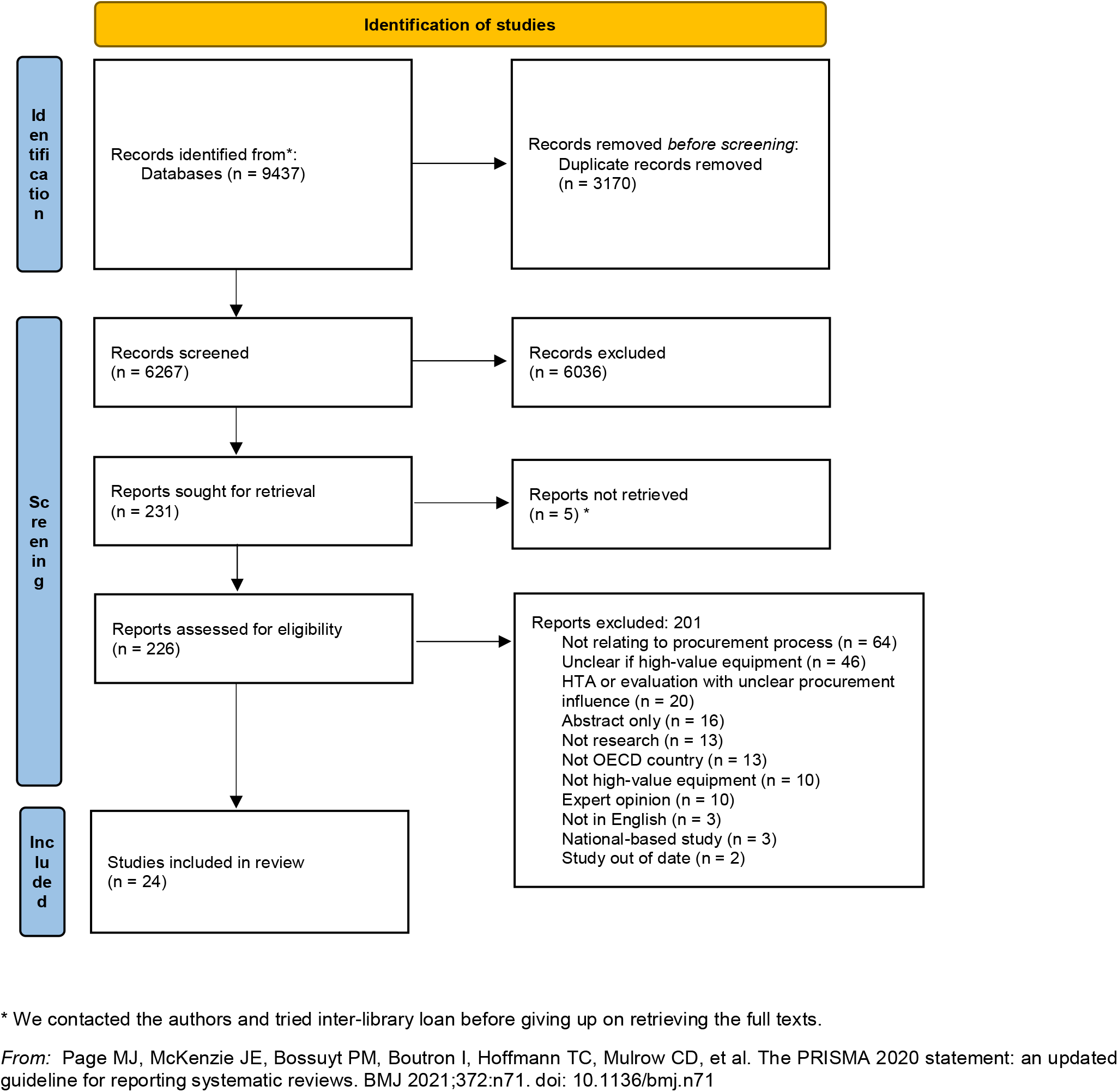
PRISMA flowchart

### Data extraction

We designed and tested the data extraction form in a spreadsheet shared via Google Sheets to enter: year in which the study was published, country in which the study took place, and number of hospitals included in the study, type of high-cost equipment that is the subject of the study (if specified), purchasing process, approach or method outlined in the study (‘intervention’), outcomes, lessons and/or recommendations emerging from the study, research method adopted in the study, limitations of the study as reported by the study authors. One reviewer extracted the information from each study, and the work was double-checked and, if necessary, completed by another reviewer. Any questions were discussed in the bi-weekly meetings.

### Data synthesis

We summarised the information from the literature in tables and lists. Because of heterogeneity of study designs across the small number of included studies, we did not conduct any quality assessment of the included studies; however, we reported the limitations listed by the researchers for their study.

### Protocol registration

This review was registered in Open Science Framework.[24]

## RESULTS

Out of an initial 9437 retrieved records, 24 studies were selected for inclusion (shown in Table 1). These included research articles (n=21), PhD/Masters theses (n=2), and one book chapter. Countries in which the hospitals were based for these studies were USA (n=10), UK (n=7), Italy (n=2), Mexico (n=2), Canada (n=2), and one from Australia, Greece, Switzerland, Germany, Netherlands, and Scotland, including cross-country comparisons. Most studies were conducted in one hospital, with a few reporting work across 2-44 hospitals. The types of equipment that were the focus of these studies ranged from orthopaedic implants, to diagnostic lab equipment, and larger investments such as MRI scanners and surgical robots. We identified a diversity of disciplines represented by the journals where these studies were published, reflecting the diversity in how the subject of purchasing high-cost medical equipment is addressed in academic work. Study types included descriptions of processes taking place within or across hospitals (n=14), which had no formal evaluations but three of which reported cost savings; empirical studies in which hospital records or participant data were analysed (n=8), and evaluations or pilots of proposed purchasing processes (n=2).

**Table 1.** Full list of included studies

| Study name | Type of article | Journal | Year | Country | Setting | Device/ Equipment | Main aim of paper | Research methods | Intervention/Approach | Lessons/Outcome | Limitations |
| --- | --- | --- | --- | --- | --- | --- | --- | --- | --- | --- | --- |
| Callea et al. (2017) [25] | Journal article | Social science & medicine | 2017 | Italy | 44 hospitals | Devices for interventional cardiology, interventional neurology, neuro-surgery, and orthopedics (distinguishing between "costly" and "inexpensive" devices) | To investigate the combined effect of various health technology assessment (HTA) governance models and procurement practices on the two steps of the medical device purchasing process (i.e., selecting the product and setting the unit price). | **Empirical study (using hospital records):** Existing survey data, document and literature review, model calculations to investigate effects | Use of regional HTA and/or hospital-based HTA functions; arrangements for centralised procurement | Regional HTA increases the probability of purchasing the costliest devices, whereas hospital-based HTA functions more like a cost-containment unit. Centralized regional procurement reports savings averaged 13.4% for most expensive products. Hospitals located in regions with active regional HTA programs pay higher prices for the same device (9.8% for costly devices). Teaching hospitals pay higher unit prices than non-teaching hospitals for costly products (34.3%). Compared with independent trusts (public hospital groups), research institutes pay 18.1% less on average for costly devices. | Devices are "neither costly nor inexpensive per se" because the definition relies not on a reference price but rather on the actual unit price paid by the hospitals in the sample. Sample size is only 18% of Italian hospitals. Study assumes costliest device is most innovative which is contested. |
| Eagle et al. (2002) [26] | Journal article | The American Journal of Managed Care | 2002 | USA | 1 hospital | Defibrillators, pacemakers, coronary stents, and coronary balloon catheters | To assess the magnitude of savings and develop concepts for "best strategies" in reducing costs in the purchasing of high-technology, high-cost materials used in coronary interventions and electrophysio logic treatments. | **Description of process (with reported cost savings):** Case study reporting on experience | Iterative negotiation following a broad request for proposal sent to a diverse group of vending organizations in high-technology areas of cardiology. Product costs and volume usage were assessed before and after the process to estimate annualized cost reduction achieved. Collaborative consensus among physicians, administration, materials management, purchasing, and vendors. | Aggressive, collaborative, fair, and competitive bidding for high-cost products used for coronary interventions and electrophysiologic treatments leads to substantial cost savings and can promote provider-industry partnerships that further enhance product use, provision, and tracking. | None listed |
| Greenwood et al. (2014) [27] | Journal article | Journal of Clinical Engineering | 2014 | Canada | 1 hospital | Capital Equipment (examples given are: table, examination; scanner, ultrasonic, bladder) | To examine the effect of a clinical engineering role change (from equipment maintenance to health technology management) | **Description of process (with reported cost savings):** Case study using experience and data from the previous three 5-year clinical capital equipment plans were collected and analysed. | Development of in-house clinical engineering expertise who develops Risk Ranking System and Long-range technology plan: (1) a theoretical replacement plan, (2) an emerging technology plan, and (3) a fleet equipment plan | Developing in-house clinical engineering (CE) expertise enables the facility to keep its capital equipment current and keep clinician acceptance high by maintaining a fair and methodical process. Hospital has made its clinical environment safer through the use of planning tools such as fleet management, equipment standardization, and a balanced request scoring system while keeping within its long-range capital equipment budgetary limits. The average age of clinical equipment has dropped substantially to just over 5 years as of the 2011 plan. Annual contingency fund expense for clinical capital equipment no longer absorbs between 15% and 25% of the overall CE budget. It has now been fixed at the relatively small amount of 5% of the overall budget, and this threshold has been reached in only 1 of the last 5 fiscal years. . | None listed. |
| Haas et al. (2017) [28] | Journal article | The Journal of arthroplasty | 2017 | USA | 27 hospitals | Prosthetic implants | To determine the drivers of variation in prosthetic implant purchase prices for primary total knee and hip arthroplasties (total knee arthroplasty (TKA) and total hip arthroplasty (THA), respectively) across providers. | **Empirical study (using hospital records):** Multivariate linear regressions to identify which variables had greatest influence on purchase price | Use of a hospital physician committee for implant vendor selection and negotiation | The use of a hospital-physician committee was associated with lower purchase prices relative to the hospitals where the physicians selected which vendors to use and the hospital separately negotiated prices with those vendors. | Small, non-randomised sample; retrospective observational study with no longitudinal data; did not assess whether hospitals changed approach during the study year; used self-reported data; not able to examine details of price variations |
| Haselkorn et al (2007) | Journal article | American Journal | 2007 | USA | 27 hospitals | Unspecified | To assess the structure, | **Empirical study (using** | Technology planning and | Having an organizational culture ready and committed to a well thought out, structured | None listed |
| [29] |  | of Medical Quality |  |  |  |  | processes, and cultural support behind hospital committees for new technology planning and approval. | participants): Survey (n=35 responses from 27 organisations ) | approval process (described as well-organised, consistent, standardised/centralised process, and with a committee with authority to give direct approval of new purchases) | approach to technology planning and assessment is a crucial component for success |  |
| Kuper et al. (2011) [30] | Journal article | BMJ | 2011 | UK | 3 hospitals | Oesophageal Doppler cardiac output monitor for fluid administration | To identify barriers to procurement and implementation of oesophageal Doppler monitoring | **Evaluation of process (across hospitals):** Comparative before (retrospectively available data from matched controls)/after (prospectively collected data from patients) study for patients' outcome data; qualitative data from survey of anaesthetists and meetings | A campaign for adopting technology in major surgical specialties explored clinical and managerial barriers throughout the procurement and implementation process. A business case was prepared by each team with support from NHS Technology Adoption Centre, allowing senior management to overcome the unequal spread of costs versus benefits. A survey of anaesthetists revealed concerns about familiarity with the device, which we dealt with by clinicians volunteering to "champion" the technique, supported by standard training provided by the manufacturer. Team encouraged appropriate use of the technology by collecting intraoperative patient related data and postoperative patient outcomes and by giving regular, timely feedback. | Managerial barriers consisted of silo budgeting, difficulties with preparing a business case, and fears about uncontrolled implementation. By collecting outcome data, we convinced senior managers to support and sustain investment. Clinical barriers consisted mainly of scepticism regarding clinical effectiveness and worries about training. Clinicians "championing" the technology took on responsibility for data collection, education, advocacy, and spanning boundaries. The project generated a web based guide to provide tools and resources to support implementation. Patient outcomes improved after managerial and clinical barriers to implementation were identified and overcome | Non-randomised "before and after" project. Despite matching for specialty and severity of operation, the control and implementation groups had differences in age and physical status scores. Results could have been confounded by other changes occurring over the same time period. At one site, in elective colorectal surgery only, a multidisciplinary enhanced recovery programme was introduced and may have contributed to the observed improvement. Any implementation study of this type is vulnerable to a Hawthorne effect, whereby performance improves as a result of close observation. |
| Langenburg et al. (2003) [31] | Journal article | Pediatric Endosurgery & Innovative Techniques | 2003 | USA | 1 hospital | Surgical robotics | To describe experiences in developing and implementing a program for computer-assisted, robot-enhanced surgery | **Description of process:** Case study based on experience | Defined a core group of individuals who shared vision: pediatric surgeons, our institutional research director, a biomedical engineer and physicist, and hospital chief executive officer. Partnership developed to continue research and development of equipment and surgical techniques. Developed short-term and long-term educational, research, and business plans; shared with hospital administration and hospital board of trustees to garner support. The staff of the hospital development office was also involved in generating financial support. | Institutional and private donor support has allowed implementation of a robotic minimally invasive surgical suite in operating room and in research building. Within one year of embarking on program the team performed our first robot-assisted minimally invasive surgery on a patient. Many of pediatric subspecialty colleagues have been utilizing suites for procedure development in their areas of interest. The key elements in developing a new program are to define a core group of committed individuals, define your vision, create corporate partners, and garner financial support with a sound educational, research, and business plan. | None listed |
| Larios et al. (2000) [32] | Journal article | Technology and Health Care | 2000 | Greece | 1 hospital | Microbiology equipment such as blood analysers and medical imaging | To streamline the management process related to procurement | **Evaluation of process (within hospital):** Process model | Proposing a procurement process for new hospital sites or expanding sites using a | The success criteria of the proposed process are time-cycle and efficiency gains in the biomedical equipment procurement procedure, Consistency gains and Information Integration, Knowledge Re-use, and shifting the core of the decision-maker's | None listed |
|  |  |  |  |  |  | modalities such as Computer Tomography(CT), Magnetic Resonance Imaging (MRI), Ultrasound and typical X-Ray equipment | to increase efficiency using a Management Information System (Biomedical-equipment Information System=BIS) | development; pilot test conducted to measure time cycle of procurement process | management information system: Addressing the tasks of: a) defining appropriate biomedical equipment specifications; and b) supporting the selection of the best bids among a huge-range of alternatives, on the basis of quality, cost and time-efficiency of the process. The proposed re-designed process was evaluated during the assessment of bids during the equipment purchasing process of the Micro-biology and Radiology Departments of a large hospital complex in Athens, Greece, as a pilot application. This paper proposes a streamlined decision-making process, addressing the tasks of: a) defining appropriate biomedical equipment specifications; and b) supporting the selection of the best bids among a huge-range of alternatives, on the basis of quality, cost and time-efficiency of the process. | work towards operations that are of more judgmental than data-handling nature. Time-cycle of the Biomedical-equipment Procurement Process has been reduced from an average of 154 days to an average of 92.5 days. |  |
| Lindgreen et al. (2009) [33] | Journal article | Journal of Business Ethics | 2009 | Netherlands | 7 hospitals & 1 private center | MRI scanning equipment | To investigate how environmental and social dimensions are perceived and how it supports health technology purchasing in hospitals | Document analysis, Focus group, interviews, questionnaire | N/A | None of Philips Medical Systems's five "green focal areas" indicators are universally considered important as influences on the purchasing decisions of interviewees. All interviewees identified health and safety as an important influence. Philips Medical Systems was perceived to engage proactively in enhancing safety during usage and equipment maintenance, based on the assumption of duty of care rather than tangible evidence. Both "operator comfort" and "patient comfort" universally are perceived as important, but their influence differs because of the involvement timescale ( operators spend their entire working day scanning, whereas patients spend just a fraction of that time). The interviewees consider both "ethical production" and "ethical production at the producer's suppliers" synonymous, but even though unethical production has high media impact, only 68% of interviewees consider this indicator professionally important, though the majority consider it personally so. Only one interviewee thought product accessibility professionally important. 90% of the interviewees believe the "contribute to science" indicator is important, because they perceive it to mean that the scanner advances the science of diagnosis. The findings highlight that not all indicators can measure performance. | single-case approach; focus on the purchasing stage, patients as customer stakeholders do not appear in the study, which limits understanding of how their views about indicators such as safety and comfort might influence the opinions of the decision makers and thus prevents are commendation about the desirability and practicability of targeting marketing effort to them. Study relies on historical information and interviewees' recall; real-time data collection could identify transitory influences on stakeholder's views, and longitudinal research might distinguish how these influences have affected company policy |
| Li et al. (2015) [34] | Journal article | Journal of Long-Term Effects of Medical Implants | 2015 | USA, Canada, Scotland | 26 hospitals | Orthopedic Implants | To determine the factors that affect purchasing decisions related to osteoarthritis | Empirical study (using participants): Qualitative Electronic Survey | N/A | Items related to clinical evidence and cost effectiveness had a greater influence than those related to a specific individual's personal preference in the process of making purchasing decisions, whether it was the administrator, surgeon, or patient. However, surgeon preference did have a higher average ranking compared to device<br>cost reassuring that patients are receiving the most clinically effective care and that the type of treatment that they receive is not heavily influenced by costs. The most important considerations for adopting new technology were whether there was sufficient evidence in the literature, followed by thoughts of key opinion leaders, and cost of intervention/device. | Canadian hospitals were underrepresented. Low response rate. Sample was more representative of smaller hospitals serving smaller populations and with a lower number of orthopedic surgeons on<br>staff. The authors may consider restructuring our survey in order to make it simpler to complete, yet capture all of the same information and hopefully encourage more participants to respond. |
| Licona et al. (2009) [35] | Journal article | International Journal of Technology Assessment in Health Care | 2009 | Mexico | 1 hospital | CT scanner | To demonstrate the experience of a managed network of professionals inputting into equipment management in one institution | **Description of process:**<br>Case study reporting on experience | Involvement of a multidisciplinary group (drawn from researchers, undergraduate and graduate students in fields that range from architecture to civil and biomedical engineering) to deal with large and complex issues within the field of hospital engineering. Steps involved specifically in the equipment planning phase include: assessing availability of similar equipment at locations in the vicinity; cost-effectiveness planning; incorporation of data on equipment availability at the state-wide level combined with morbidity and mortality figures, incorporation of information regarding "plant" installations including electrical, hydraulic, and telecommunications. Specifically for the case of the CT scanner purchase: The BME branch of this group analyzed the bidding procedures, the contracts and asked several questions that needed to be answered before the formalization of the reception could be signed. | During this study, several anomalies were discovered: The equipment being bought was constructed by one of the three major vendors of imaging equipment worldwide. However, they did not participate in the bidding process. A local company won the bid and then proceeded to subcontract the equipment from the major vendor. The questions arose as to who was installing the equipment, because it appeared that the major vendor was providing the technicians, which was a breach of contract (bid-winning companies should provide training and do installations themselves). A second question arose regarding the existence of replacement parts within the winning company's warehouses, and finally, there was a major question posed as to the adequacy of the equipment being bought (sixty-four-slice CT specially built for cardiac studies) for a general hospital with no cardiac specialties, as well as the elevated sale price (as much as a magnetic resonance imaging scanner). The hospital took these results in hand and acted in accordance to its administrative procedures to correct the anomalies | None listed |
| Lingg et al. (2016) [36] | Journal article | BMC Health Services Research | 2016 | Mexico, Germany, Switzerland, UK | N/A representativ es across countries and settings | Orthopaedic devices (high-risk) | To better understand the impact of procurement on clinical procedures and outcomes | **Empirical study (using participants):** 59 in-depth interviews with stakeholders from Mexico, Switzerland, Germany, and UK: orthopaedic specialists, government officials, other experts, and social security system managers or administrators | Involvement of orthopaedic specialists in procurement process, and use of post market surveillance data to inform decision-making | Procurement processes for orthopaedic HRMDs may have an impact on clinical practice and outcomes. Three areas of deficiency were identified: 1) HRMD regulations based on insufficiently robust clinical evidence (mainly noted by European countries); 2) Follow-up on Health Technology Assessments is inadequate (noted by Mexico) and methodology not always good enough (noted by European countries); and, 3) Lowest-acquisition price often guides procurement decisions and thus may not align with needs of clinical procedures (noted by Mexico and some European countries) | Micro level stakeholder (patients or representatives from rehabilitation centres) not included in study. |
| Madhlambudzi and Papanagnou (2019) [37] | Journal article | International Journal of Healthcare Technology | 2019 | UK | 2 hospitals | Diagnostic equipment | To describe analysis of decision-making processes when the public | **Description of process:** Case studies and semi-structured interviews (n=121, | N/A | NHS hospitals fail to identify key stakeholders resulting in possible delays and conflicts. Throughout our research, it was ascertained that NHS hospitals do not tend to apply stakeholder analysis as a part of their project planning process. This has in some cases resulted in leaving out key | None listed |
|  |  | gy and Manage ment |  |  |  |  | hospitals purchase diagnostic equipment and it discovers how the hospitals use stakeholder identification and salience during the purchase of diagnostic equipment | narratives of people involved in decision making on outsourcing laboratory diagnostic equipment), document analysis |  | stakeholders and thereby bringing about conflict and delays in the process. NHS hospitals are bound by strict guidelines in their procurement processes to avoid bias and ensure competition among potential suppliers and get the best deal. Technical personnel, however, came up with some valid reasons why it would be more suitable to upgrade the present equipment than to undertake radical adjustments or changes. It is, therefore, important that at any stage of the process the weight of the stakeholders should be considered in deciding whether their input is acceptable or not. |  |
| McCue (2011) [38] | Journal article | Health Care Management Reviews | 2011 | USA | Short-term acute hospitals in state of California (number unspecified) | Unspecified (Capital expenditures of equipment included CTscanners, MRIs, picture archiving and communication systems, and surgical systems) | To identify the market, organisational, and financial factors associated with capital expenditure projects (of which capital medical equipment was one category) | Empirical study (using hospital records): Secondary data analysis: association study using ordinary least squares regression analysis on retrospectively collected hospital capital expenditure data from 2002 to 2007 | N/A | Hospitals located in urban markets with greater share of the market had a greater number of medical equipment purchases per hospital. Hospitals with greater market share had a greater number of medical equipment purchases per hospital. The positive coefficient for hospitals with over 350 staffed beds suggests that these facilities had a greater number of medical equipment purchases per hospital, whereas negative coefficient for hospitals with less than 100 staffed beds had fewer number of medical equipment purchases per hospital. The positive coefficient for system affiliation indicates that hospitals owned by large systems had a greater number of medical equipment purchases per hospital. Hospitals with greater liquidity had a greater number of medical equipment purchases per hospital. hospitals with an aging plant and equipment had fewer number of medical equipment purchases per hospitals. Hospitals serving a greater percentage of government payers had fewer medical equipment purchases. Teaching hospitals had greater number of medical equipment purchases per hospital. Investor-owned hospitals had fewer medical equipment purchases. | The primary limitation of this study is that the findings can only be generalized to the state of California. |
| Mitchell et al. | Journal | Internati | 2010 | USA | 1 hospital in | Cardiac | To describe | Description | Integration of local | Hospital-based HTA using local data can fill | While analyses were done |
| (2010) [39] | article | onal Journal of Technology Assessment in Health Care |  |  | 1st case; 3 hospitals in 2nd case. | catheterization lab; ICU telemedicine services | two evidence reports from our hospital-based HTA center which required the integration of local data. Both cases illustrate how local evidence can be used at the institutional level to support the quality, safety, and cost-effectiveness of patient care. | **of process:** Two case studies (one using qualitative and one using quantitative data); 1st Case: equipment service records, and interviews with physicians, technicians, and administrative staff. 2nd Case: systematic review of effectiveness of service, the hospital's administrative and claims databases (including Mortality and Length of stay) | qualitative and quantitative data into hospital-based HTA to select a new technology or inform a decision on whether to continue services. | gaps in the published evidence, and also improve the generalizability of evidence to the local setting. To take advantage of local evidence, health systems should encourage the development of hospital-based HTA centers, seek out local preference data, and maintain databases of patient outcomes and utilization of services. The use of local evidence to support institutional decision making can also reduce problems of external validity. In both case studies, important differences among the hospitals within health system was found. These differences affect the prioritization of different attributes of a technology, and could result in different conclusions being drawn about how the technology should be used at each hospital, even within the same healthcare network; the experience and expertise of local clinicians should be respected when making decisions at the hospital or health network level (it helps decision makers understand possible differences in local patient populations or in processes of care that may affect the cost or effectiveness of the technology, and it promotes "buy-in" from the clinicians who must implement the decision). | in retrospect (Data have to have been collected and available for analysis), the research could not control variables such as changes in staffing or new infection control policies. In analysis of ICU outcomes, the study lacked APACHE scores for ICU patients before the introduction of telemedicine coverage, so the ability to control for patient acuity was limited. The available claims information did not include enough detail to ascertain whether possible lapses in care happened in the ICU or elsewhere. While there was no such problem with availability for the survey data used in cardiac imaging decision, gathering that data required considerable fieldwork. |
| Mosessian (2016) [40] | PhD thesis | NA | 2016 | USA | Multiple hospitals (unspecified) | Orthopaedic implants | To examine the extent to which Value Based Purchasing is being used to purchase implanted orthopaedic medical devices, and the decision-making processes | **Description of process:** A survey tool was developed (with input from a focus group with 10 professionals) and responses obtained from two groups of | Use of Value-based committee: physicians and surgeons make decisions, hospital administrator makes decision, bundles corporate purchase agreements, request for proposals issued, group purchasing organisations. | Results include: (1) the two most important decision-making attributes for both groups were quality of care and cost-containment. (2) most health care settings now use decision-making systems more amenable to value-based purchasing than previous ad-hoc decisions driven by surgeons, (3) decisions are commonly, but not universally, made by committees with representation from surgeons, administrators and often others, who work together to choose implants, and that (4) their processes are still mostly based on information derived from the clinical experience of clinicians and | Data based on USA hospitals only; reimbursement entities, patients nor regulators' views not included; general limitations of survey responses noted. |
|  |  |  |  |  |  |  | that are being implemented to support those acquisitions. | stakeholders, hospital executives (n=29) and orthopedic surgeons (n=40) | Intervention specifically studied: value based purchasing and knowledge of procurement officers use (rather than HTAs) | local knowledge of procurement officers, with less influence from more formalized health technology assessments. |  |
| Nisbet et al. (2001) [41] | Journal article | The British Journal of Radiology | 2001 | UK | 1 hospital | Radiotherapy equipment | To describe financial factors affecting decision to purchase or lease radiotherapy equipment in one hospital and to describe technical consideration to be taken into account | **Description of process:** Case study. Financial analysis (over 10 years to correspond with the assumed economic lifetime of the equipment) and Operating Lease Test | Overview of the procurement process, including a summary of the advantages and disadvantages of leasing, with the figures from the financial analysis; a detailed description is given of the technical considerations to be taken into account in the financial analysis and negotiation of any lease contract. Comparison of leasing as defined in the Statement of Standard Accounting Practice 21 (SSAP21) and purchase. | It is essential that technical staff are involved in the discussion and detailed negotiations on the content of the lease, and ideally the financial aspects of these considerations should be taken into account during the financial analysis of purchase vs lease. | Larger centres with a rolling programme of replacement equipment would expect to keep up to date with technological advances, and the conclusion reached for this hospital may not apply. |
| Obremskey et al. 2012 (Vanderbilt case) [42] | Journal article | Clinical Orthopaedics and Related Research | 2012 (2008 start of intervention) | USA | 1 academic medical centre | VANDERBILT Case: Surgical Implants (Physician Preference Items): Surgical endomechanical stapling devices, orthopaedic joint arthroplasty, spine internal | To describe the challenges, implementation, and outcomes of cost reduction and product stabilization of a value-based process for | **Description of process (with reported cost savings):** Case study | Vanderbilt case: Implementation (2008) of a physician-driven Facility-based Technology Assessment Committee (=Medical Economic Outcome Committee) that standardized and utilized evidence- | Utilizing this physician-driven committee, we provided access to new products, standardized some products, decreased costs of physician preference items 11% to 26% across service lines, and achieved savings of greater than \$8 million per year. The implementation of a facility-based technology assessment committee that critically evaluates new technology can decrease hospital costs on implants and standardize some product lines. | VANDERBILT: First, the study describes the experience of only one institution. Each institution has its own challenges in physician alignment, history, and culture. Each institution's process will be unique to its individual characteristics. Second, the institution is an academic setting with closely aligned faculty and |
|  |  |  |  |  |  | fixation, trauma internal fixation, cardiac rhythm management implants, drug-eluting stents, and cardiac valve implants. In Table: Endomechanical, Total joints, Cardiac rhythm management, Drug-eluting stents, Spine implants, Interventional cardiology, Cardiac surgery, Trauma, Abdominal mesh. 2013 report: + Closure Devices, Transcription, Oral Care, and Reference Lab Phase I. | purchasing medical devices at a major academic medical center. |  | based, clinically sound, and financially responsible methods for introducing or consolidating new supplies, devices, and technology for patient care. This committee worked with institutional finance and administrative leaders to accomplish its goals. |  | hospital. Academic practices that are not directly affiliated with the hospital and community hospital with community-based surgeons will have to establish a mechanism to partner with each other for mutual benefit. Third, the institution established the committee a short time ago, and long-term effects of the process cannot be described. Finally, while other institutions could reproduce this process, it will not guarantee the reproducibility of the effects of this study. Each institution will need to develop and modify the described process to fit the culture, history, and geography of their situation. |
| Olson et al. (2013): Cases: Vanderbilt and Duke [43] | Journal article | Clinical Orthopaedics and Related Research | 2013 (Intervention since 2008 and 2010) | USA | 2 academic Medical Centers | DUKE: Endo-Mechanical, Total Joints, Cardiac Rhythm Management, Drug Eluting Stents, Spine Implants* (Hardware Only), Trauma, MESH, Heart Valves Rings, Nerve Stimulation, Kypho-Vertebral | To describe physician-led processes for introduction of new surgical products and technologies; and to inform physicians of potential cost savings of physician-led product contract negotiations | Description of process (with reported cost savings): Case studies (2) | Duke case: Implementation (2010) of Medical Staff Committee with a charge to evaluate Equipment, Devices, and Information Technology (EDIT) to be brought into the operating room (OR) | A collaborative arrangement should address three objectives in which hospitals must find ways to meet three objectives: (1) collaborate with medical staff leadership to provide surgeons with feedback regarding the financial impact of their implant selection on the cost of an episode of care; (2) ensure that medical staff leadership has an effective means of communication with hospital administration regarding the medical evidence supporting the use of newer, more expensive technologies or implants to benefit patient care; and (3) both the hospital and physicians need a system that allows tracking of the impact of efforts to manage implant use. There are | See Obrebsky et al. 2012 + First there is very little peer-reviewed research and literature in this area. Second, the experiences in academic centers may not be applicable to other environments. Third, to achieve physician participation in these programs, some higher form of alignment between physicians and hospital or the health system must be in place. Fourth, we have very little |
|  |  |  |  |  |  | Plasty, Negative wound pressure, EP Catheters and Accessories, Bare Metal Stents, Duke University Hospital System total. VANDERBILT: endo-mechanical, total joints, cardiac rhythm management, drug eluting stents, spine implants, closure devices, interventional cardiology, cardiac surgery, transplantation, trauma, MESH, oral care, reference lab phase I. | and approval of new technology. |  |  | potential disadvantages in setting up a physician-led system as well. For physicians leading such efforts, a substantial amount of time may be required. The value for hospital systems from these programs is centered around cost savings, whereas the value for surgeons is centered around access to technology and products required for cutting-edge medical care. Thoughtful communication to each of these key groups of stakeholders is necessary to ensure the successful work of the program is shared to each group. | published peer-reviewed data on cost savings. Such data will need to be accumulated in the future in a form that can be subject to peer-reviewed publication. |
| Pandit et al. (2011) [44] | Journal article | Anaesthesia | 2011 | UK | N/A | airway management devices | To establish a process to create appropriate level of evidence to inform purchasing decisions within hospitals (in UK) with a working party (Airway Device Evaluation Project Team) | **Description of process:** Case study of process developed to support adoption | Difficult Airway society working party advises on how to set up design of a trial appropriate specifically for airway devices and guides hospital in implementation of this trial together with company (who sponsors it); results published for other hospitals and results in final purchase | NA - does not report on implementation of proposed procurement process | ("Weaknesses of strategy") ADEPT's decision to leave many judgements to individual discretion was a pragmatic one, and arguably, there is not enough dictated from the centre. Some trusts may continue to ignore anaesthetic opinion, prioritising instead the financial consideration. Some manufacturers may try to use a non-evidence-based approach to marketing their products. |
| Saaid et al. | Journal | American | 2011 | Austr | 4 hospitals | Unspecified | To examine | Empirical | Use of business | Decision making processes were described | None listed |
| (2011) [45] | article | Medical Journal | (Study in 2010) | alia |  |  | the decision-making processes for acquiring new health technologies in selected hospitals, guided by approaches from a decision-making model and a mini-Health Technology Assessment (HTA) model | **study (using hospital records and participants):**<br>Two Studies:<br>1. A multiple case study method using convenience sampling: Document analysis (mini-HTA checklist as a benchmark) and 2. Qualitative: In-depth, face-to-face interviews via content and thematic analysis | strategy and cost effectiveness analyses. | as informal in not-for-profit private hospitals and as formal in public hospitals. At the public hospital, HTA is a requirement for new health technology decision making. Decisions in not-for-profit private hospitals were driven by business strategy and the cost effectiveness of the technologies. In the public hospital, the main factors were safety and clinical effectiveness although budget also has some impact. The costs of the new technologies determine the complexity of the decision processes. In the public hospital, the ethics and legality of the technologies also affect the decisions. The impact of HTA as a support tool for decision makers at institutional level is still relatively minimal. Decision makers in both types of hospitals were unclear about HTA and its agencies. They also were not aware of mini-HTA, even though they were searching for a suitable support tool for decision making. The respondents stated that an open and innovative organisational culture was critical as a facilitator for the adoption of new health technologies, whereas limited resources and space were seen as major barriers. Respondents did not view human resources as a factor, because staff can be trained and up-skilled. Participants from the Public hospital believed that bureaucracy is also an important barrier to the introduction of new technologies. Resistance to change among the staff is another barrier. In terms of future improvement, 90% of the decision makers in the Private hospitals believe that the decision making process should be more structured, because structured processes ensure that the decisions are supported by facts and will reduce unfairness and prejudiced responses. Participants also spoke about timely information, they want the information be there when they need it, because the technologies are rapidly change and after one or two years there will undoubtedly be a newer technology available. Participants also believe it would be valuable if they could get information on new technology from an independent body, such as HTA agencies. The participants from public hospitals suggested that the product review committee members in their hospital should have more variation in membership so as to include representatives from doctors, nurses, pharmacies, and administrators, and not just from nurses. |  |
| Satta et al. (2019) [46] | Book chapter | Clinical Engineering Handbook (Second Edition) | 2019 | Italy | 1 hospital | ophthalmic surgery femtosecond laser | To describe a tender of ophthalmic equipment | Description of process: case study based on experience | To test a procedure for regional public tender purchase (ESTAR) including: accessories, consumables needed for sustained use, quantitative/financial evaluation (all included in the contract for true costing, which includes number of interfaces with technicians expressed in days, and limitations set in contract for locking prices over 5 years). User "trial" performed for 10months to test each option in real-life settings. | ESTAR tender procedure gave an excellent result in terms of quality of equipment and awarded prices but the total time to achieve the result is quite long. (±4 years) | During the installation, emerged technical problems could probably be addressed during the tender design phase. Furthermore, the aspects related to the data flow would have the deserved deeper analysis already from the drafting of the specifications and then also during the assessment. |
| Verma & Peacock (2014) [47] | Journal article | Ultrasound | 2014 | UK | 1 hospital | Ultrasound imaging | To describe the management structures concerning ultrasound equipment in hospital. | Description of process: Case study based on experience | Use of medical equipment management group | Medical equipment management group created successes: 1) oversight of ultrasound equipment improves handling financial implications and plan yearly expenditure 2) consolidating equipment from one manufacturer in a department improves procedures 3) redistributing equipment within hospital prevents unnecessary buying 4) buying with research funding; maintenance costs after grand period taken into account | None listed |
| Wong (2007)<br>[48] | Master<br>thesis | NA | 2007 | UK | 2 hospitals | Case 2 most<br>relevant: x-ray<br>equipment | To generate a<br>detailed<br>understandin<br>g of the<br>relationship<br>between the<br>risks which<br>the private<br>sectors bear<br>and the<br>returns they<br>actually earn,<br>to highlight<br>how risks are<br>allocated<br>appropriately<br>with the<br>stage of the<br>procurement<br>process, and,<br>to identify<br>how the<br>current risk<br>management<br>model control<br>and manage<br>Public<br>Finance<br>Initiative (PFI)<br>project risks | **Description**<br>**of process:**<br>Two case<br>studies:<br>interviews,<br>questionnaire<br>, document<br>analysis | Use of PFI<br>procurement | Risks in PFI contracts are appropriately<br>transferred and mitigated under the current<br>risk management system in technology and<br>equipment management NHS projects. The<br>transfer of technology and obsolescence<br>risks to the private sector is fundamental to<br>the delivery of Value For Money (VFM) in<br>PFI procurement in health sector. PFI<br>procurement in hospital projects results in a<br>more structured approach to operating,<br>maintaining and replacing medical<br>equipment assets. | None listed |

Although excluded in our own review during full-text filtering, we had identified 20 studies that combined HB-HTA or other assessment methods with decision criteria directed towards a purchasing decision, which we had to exclude because of their lack of clarity on whether these methods had direct influence on the purchasing process or final decision itself within a hospital context. Examples include Jurickova et al 2014 using value-enginering and multicriteria methods,[58] Girginer et al 2008 using analytical hierarchy methods,[59] and Hospodková et al 2019 using hospital-based HTA.[60]

### Key findings from studies

The two most prominent elements of purchasing processes identified across most of the included studies were (a) the roles of various stakeholders involved, and (b) the approaches to balancing technical, financial and clinical requirements.

#### Stakeholders and teams involved

Table 2 shows the involvement of roles in the procurement process as mentioned in the included studies, representing a combination of roles either involved in the studies themselves, and in the project teams observed in the studies. The studies reviewed were specific and emphatic about the importance of stakeholders as part of the decision-making process, specifying who exactly should be involved and how. Two stakeholder groups in particular were emphasised: clinicians and the clinical engineers, sometimes explicitly as the sole focus of the study, and at other times mentioned implicitly as part of the process. Greenwood et al 2014 reported on how the role of the clinical engineer in a children’s hospital in Canada progressed from a primary responsibility in equipment maintenance to health technology management more generally.[27] Madhlambudzi & Papanagnou(2019) studied the involvement and salience of several stakeholders in purchasing of diagnostic equipment and found that hospitals fail to identify key stakeholders resulting in possible delays and conflicts.[37] Haas et al. (2017) concluded that a hospital committee resulted in lower purchasing prices than when physicians selected vendors directly in a study of the selection of prosthetic implants.[28] However, committees are not flawless; Licona et al (2009) described a case study to demonstrate involvement of an interdisciplinary network of professionals in health technology management: despite the involved network several anomalies were identified such as uncertainty of who would install equipment after a bidding process.[35]

**Table 2:**
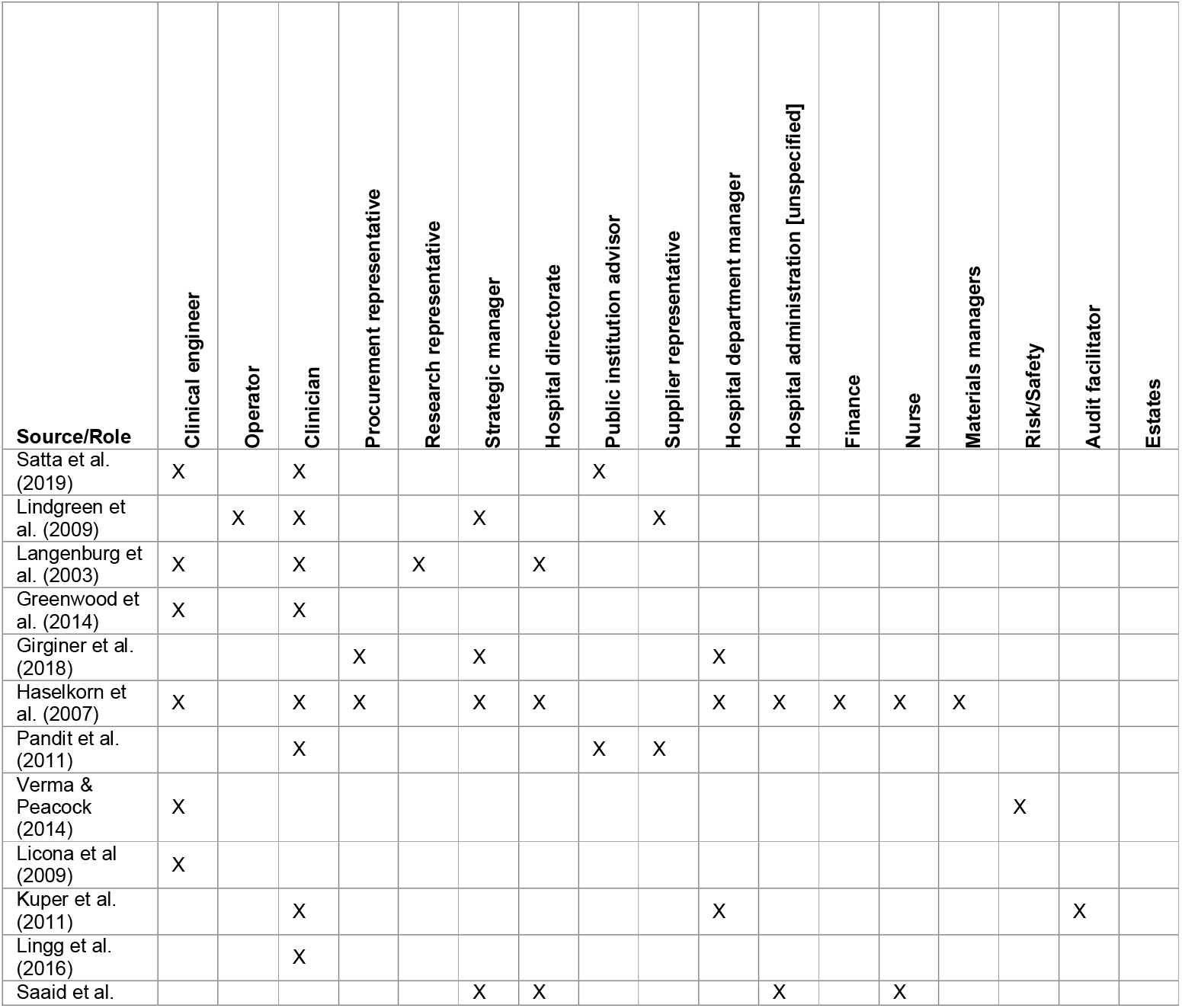
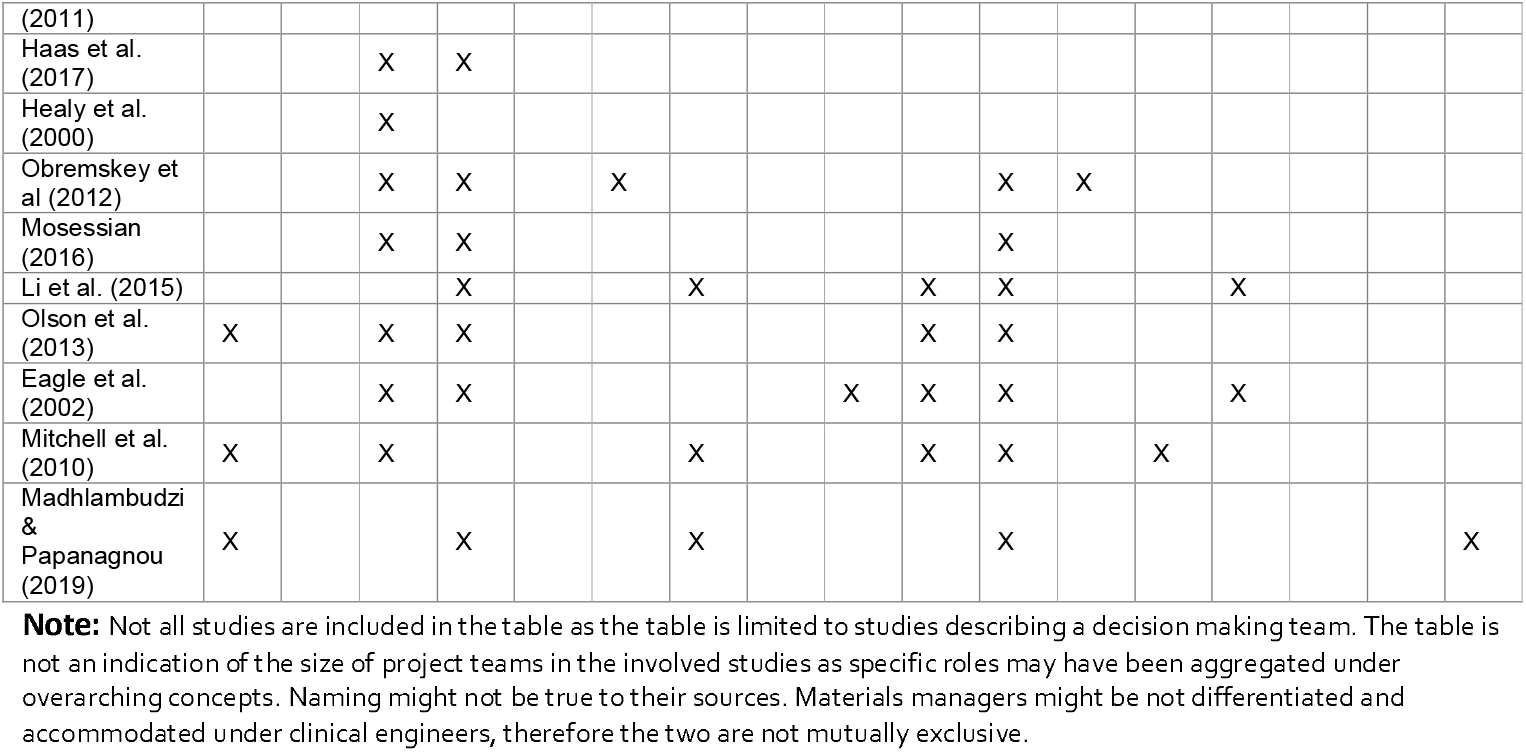
Stakeholders involved in purchasing processes as identified in the studies

Although not always the primary focus of the study, some made explicit that some form of approach that unifies how various purchasing stakeholders come together is important: Langenburg et al 2003, for instance, describe their new process as developing a ‘vision’ with paediatric surgeons, research director, a biomedical engineer and a physicist and the hospital chief executive officer, to collaborative (with industry partners) develop a short- and long-term education, research and education plan for robotic surgery.[31] Haselkorn et al (2007) also described the importance of an organizational culture as a crucial component for success in the procurement process.[29] Regardless of it being a cultural or difference in vision, fundamental differences in purchasing projects can be identified. McCue (2011) identified differences in market, organizational and financial factors associated with capital expenditure between hospitals of different size (e.g. beds) or located in different areas (e.g. urban, rural).[38] Finally, two studies specifically elicited challenges and barriers to effective purchasing. Kuper et al (2011) identified barriers to procurement and implementation of oesophageal Doppler monitoring in three UK hospitals, noting that silo budgeting and skepticism about new products challenged investment decisions; which were overcome by ‘championing’ the technology via clinicians while providing evidence of the potential benefits of the proposed technology.[30]

#### Evaluating technical, financial, and clinical elements

In the procurement of high cost, often specialized medical equipment it is necessary to balance technical, financial and clinical factors as different interests are at stake. In essence a hospital is often a company which means in the long run it should be financially feasible, but companies with big personal interests for its clients, the patients. Continuity and quality, or safety, must be guaranteed by setting technical requirements and at the same time advanced (or novel) interventions must be continuously developed and challenged in clinical aspirations. Langenburg et al. (2003) described a program combining technical, financial, and clinical elements condensed in a training, implementation and development program for surgical robotics, and found that cooperation of surgeons, staff, and a corporate partner were key to the development of a successful new program (e.g. within one year minimally invasive surgery on a patient is performed).[12] Nisbet et al (2001) describe a process in which financial and technical considerations were taken into account to decide on whether to lease or purchase radiotherapy equipment.[41] Li et al. (2015) ranked factors that influence purchasing decisions and demonstrated that clinical evidence and cost effectiveness are more important than personal preference, regardless of the stakeholder role.[34] Another example of combining multiple disciplines in order to successfully reduce costs is implementing a value based process.[40,42,43]

In order to evaluate the clinical, technical and financial elements, more formal methods are described in some studies. Pandit et al. (2011) describe a working party set up nationally to advise on how to set up a ‘trial’ specifically for airway devices and guides hospital in implementation of this trial together with company (who sponsors it); results published for other hospitals and results in final purchase.[44] The notion of more information or ‘evidence’ to inform selection is reported in different ways. Satta el al. 2019 conducted ‘user trials’ for 10 months to test each ophthalmic surgery femtosecond laser in real-life settings before selecting a supplier.[46] Other studies reported on the role of hospital-based HTA as a means to bring evidence into decision. Mitchell et al. (2010) describe how hospital based HTA provides more reliable data to the selection process by including local data when there is too little peer-reviewed evidence.[39] According to the study by Callea et al. 2017, hospital-based HTAs turn out to serve mainly as a cost containment tool in the selection process while at the same time hospitals using this method are found to pay actually 8.3% more for the same equipment.[25]

### Additional findings

In this section we report on approaches and processes identified less frequently across the included studied. Less prominent approaches and processes identified in the studies included the need for strategic and long-term planning, streamlining management processes, varied approaches to the tendering process, and relationships with suppliers. Greenwood et al 2014 described a system in which clinical engineers adopt the role of a long-term manager for health technology using three long term planning variants (e.g. theoretical replacement, emerging technology and fleet equipment), resulting in an improvement in safety and continuation of clinician acceptance.[27] A suggestion to streamline the management process is the implementation of a management information system described by Larios et al. 2000,[32] where necessary information for specification and selection of medical equipment can be documented and it is found to improve timeliness, procedural efficiency, consistency and information integration. For the development of new programs a business plan is essential), according to two studies[29,31] and proper planning and management can result in prevention of unnecessary buying according to Verma and Peacock 2014.[47] With regards to tendering, Satta et al 2019 described a process in which stringent specifications were laid out in a tender specifications for an ophthalmic surgery femtosecond laser, but note the disadvantage that their whole process of laying such specific specifications and conducting trials took about 4 years.[46] Licona et al. (2009) describe several iterations in the specification process to avoid last minute changes, and discuss that stringent specifications may lead to the selection of products with the lowest technical and qualitative requirements.[35] In another study, less stringent tender specifications actually showed to lead to substantial cost savings: instead, an iterative negotiation process with multiple vendors after a broad request for proposals led to an aggressive form of competition with varying strategies to form a solution.[26] Finally, there appears to be a reciprocity between industry and hospitals: as clinical trials with equipment have the potential to deliver evidence of functionality for devices, healthcare and industry are incentivised to cooperate in creating and obtaining this evidence.[44]

## DISCUSSION

In this systematic review we sought to identify studies that focus on approaches to purchasing of high-cost medical equipment in hospitals, in high-income countries (using OECD countries as a proxy indicator for higher income). Given the heterogeneity of study designs considered in this review, we did not apply formal quality rating system to the studies, and did not seek to find examples of ‘best’ practices, but rather attempt to identify and describe any empirical work conducted in hospital environments focussing on purchasing processes, to characterise the nature of the studies and types of approaches or interventions reported.

### Limitations of this review

We note in our introduction that this review fulfils a gap in current academic literature, which is the evidence on empirical work conducted in hospitals for purchasing medical devices and equipment. We only partly fill this gap because our review is limited to ‘high-cost’ equipment and to high-income countries, resulting in a limited picture of the purchase of other materials, supplies and devices in hospitals in a variety of contexts. Our main reasoning for this is the very different nature of processes and financial accounting for higher cost equipment in hospitals compared to lower cost devices, consumables and other supplies, which helped give a specific focus to our study. However, we found the distinction between high- and low-cost extremely challenging and consulted expert practitioners involved in hospital purchasing to advise on an appropriate demarcation, and checked for conflicts in inclusion decisions across the review team. However, we also note that studies that did not specify whether they were dealing with high- or low-cost equipment were excluded (n=47 during full text review), although some important insights could have been drawn from these. Finally, we note that another major limitation is that investment decisions do not only account for the single price of a product, but might be creating a contract of high value through bulk purchases of lower-priced devices. Again, through consultation with our experts we concluded that these specific demarcations can vary between hospitals within and across countries, and the themes derived from our review are still helpful indications of how these processes work.

Conference papers in the field of operations management and supply chains can provide useful insights into current innovations in the field – we did include them if the full text was available for review, but had to exclude those with only abstracts available. We note that we excluded studies not written in English (about 40 studies post-2000) which might have included important lessons of practice and research conducted in various global settings. During our first exclusion step (abstract/title) we came across many articles written by professional and academic experts, with no reported empirical work, but potentially extremely useful experiences to inform future practice. As our study was limited to academic research, these were excluded but could provide the basis for a targeted review of professional practice. Finally, we defined the scope of this review to start when the need for equipment is identified. We note that this leaves out a major factor of influence to the technology management process: how the need is identified, which can influence cost containment and risk assessment further down in the procurement process.

### Limitations of the reviewed studies: the nature of ‘evidence’ in this field

The motivation for conducting this review stemmed from an initial scoping search for literature on how different disciplines and researchers approach the subject of purchasing in hospitals. We sought empirical work (broadened to include single case studies) in order to provide an overview of the current evidence base for approaches to purchasing of high-cost medical equipment in hospitals. However, only three studies included any form of evaluation of their ‘purchasing process’ intervention, including one which was a pilot study based on the model developed in the study. The majority of the studies described the purchasing process in the hospital and reported outcomes such as cost savings, but did not fully report how these outcomes were assessed. We concluded that there is not yet a solid ‘evidence base’ for how to improve the process of purchasing. Conscious that we make this conclusion for studies only of high-cost medical equipment, we propose that more research that encompasses a variety of health technologies in intramural care settings can begin to provide a more comprehensive evidence base. Despite our limited focus, however, our conclusions echo those made by previous studies. A review of non-health approaches to purchasing and supply chain management literature noted that empirical work was limited, and studies “frequently fail to assess (or describe) the robustness of their methodological approaches when linking interventions with outcomes, such as cost savings or improved performance”.[16]

Conducting strong empirical work in this domain can be challenging: the theories, frameworks and methodologies necessary to address the organisational domain of healthcare (of which purchasing is one component) need to be drawn from fields such as operations research, economics, and supply chain management, and draw on approaches such as decision theory, and systems and design approaches. This presents challenges: first, the fields of purchasing and supply chain management, for example, has in itself been criticised for the lack of strong empirical work[49] and poor quality of theoretical development and discussion, and coherence,[50] and second, the application of these approaches in real health care settings has also been limited, exemplified by a recent systematic review of application of systems approaches in healthcare.[51] A recent review on logistical parameters within international research on hospitals noted that “the international literature does not, by definition, reflect what really happens in hospitals.”[52] Generally, it has been noted that evidence-based management (if we consider procurement processes to fall under a hospital’s management) in healthcare is not yet commonplace and takes various forms.[53]

### Implications for practice: lessons learned for hospital purchasing

Despite the limitations discussed above, there are some repeating actions identified in our studies that have implications for practice. Specifically, the necessity of bringing together a skilled multidisciplinary team for large investment items is highlighted across most of the studies as the key ‘intervention’ for their purchasing process. We recognise these are not conclusions made based on evaluations, but their prominence in reporting this as a key feature merits its mention. Specifically, the role of the clinician in some form of committee or decision team is emphasised, as well as the clinical engineering team as a genuine stakeholder in the final decision. Studies conducted elsewhere on lower value equipment have also highlighted the role of the clinical engineer, and the WHO’s technical series on medical device procurement specifically mentions clinical engineers as the primary role for health technology management in hospitals.[54] But how seriously this role is taken when it comes to the final investment decision remains unknown in practice.

The second most prominent theme across the studies is the importance of balancing technical, financial and clinical requirements, specifically by using some formalised method for this assessment. This could be implemented through user trials to gather the necessary evidence on device performance, literature reviews or indeed through a formal hospital-based HTA process. However, we note from some of the other studies we came across on the emergence and progress of HB-HTA, that there is limited evidence on whether or not these processes end up influencing investment or purchasing decisions (see, for example, Gagnon 2014[55] and Almeida et al. 2019,[56] and research suggests that there has been a low to moderate use of economics frameworks or value-oriented decisions in local hospital technology decision-making.[57]

### Implications for future research

Based on the limitations and implications discussed above, we recommend where research is needed to improve the evidence base for improving medical equipment purchasing decisions in hospitals. First, the demarcation challenges identified earlier (in our case, between high- and low-cost equipment), highlight the importance of encouraging specificity in studies pertaining to any management of technology in hospitals in future research. Some studies simply mention ‘supplies’ or ‘materials’ or ‘technology’ or ‘equipment’, and are insufficient to glean best practices and to ascertain how the lessons learned from the studies can be applied in both future research and practice. Specificity can also help create other ways of investigating the processes for different types of hospital purchases: in practice, many materials and supplies tend to involve different processes simply depending on their cost (and not unit cost, but cost of the whole purchase contract). Future studies could also investigate how creating processes differentiated by risk (or patient safety or criticality) rather than cost, would affect the effectiveness of the purchasing processes in supporting clinical needs. Second, it would be worth investigating the increase in assessment and evaluation methods (such as HB-HTA and human factors engineering), and how this connects and affects the ultimate purchasing decision. Connecting HB-HTA to final hospital investments in particular has been shown to be limited, the research challenge would be to investigate why, whether and how barriers need to be overcome to enable more evidence-informed hospital purchases. Finally, we challenge the research community to increase the evaluation of interventions within hospital’s organisational domain, explore the application of theories from different disciplines (including, but not limited to, operations research, engineering design, systems theory and decision theory) in this domain, and use future empirical work to further inform the theoretical advances back into those fields.

## CONCLUSIONS

In this review, we sought to identify studies that focus on the purchasing of high-cost medical equipment in hospitals, in high-income countries. Our 24 included studies point to the importance of multidisciplinary involvement (especially clinical engineers and clinicians) in purchasing decision-making to balance technical, financial, safety and clinical aspects of device selection, and highlight the potential of increasing evidence-informed decisions using approaches such as hospital-based health technology assessments or conducting user ‘trials’ of the device in use before purchase. Our recommendations for future research is to have increased specificity in the types of materials, devices or equipment being studied and reported, given that the diversity of such purchases with and across hospitals globally means lessons learned can otherwise not be applied in practice. Echoing other scholarship on the domains of management, operations research and supply chain management, we advocate for more intervention-based and empirical work to advance the evidence base in this domain.

## Supporting information

PRISMA checklist

## Data Availability

All data produced in the present work are contained in the manuscript

## OTHER INFORMATION

## Acknowledgements

We are grateful to the expert practitioners working in hospital purchasing who provided guidance and advice during this project.

## Author contributions

FS and SHK drafted the protocol. HB, BD, AC, JE commented on the draft protocol. FS and JE piloted the title and abstract screening stage for the first 500 records. FS completed the first round of screening. SHK, HB and AC screened the Included and Maybe folders. SHK made the final decisions when disagreements continued. FS, HB, and BD extracted the data and SHK double-checked and completed the extracted data when needed. SHK and BD summarised the results and drafted the final report. All authors read, commented, revised and approved the final manuscript before submission.

## Ethics statement

This review did not involve experiments on any animal or human subjects.

## Patient and Public Involvement

This review involved studying of academic literature only and therefore the involvement of patients or the public was not applicable.

