## Appendix 1 for "Purchasing high-cost medical equipment in hospitals: A systematic review"

**Appendix 1 – Search strategies**

**Cost-Effectiveness Analysis Registry**

Search for Methods

| 1 | Procurement | 17 |
| --- | --- | --- |
| 2 | Procuring | 2 |
| 3 | Procure | 17 |
| 4 | Procured | 1 |
| 5 | Purchasing | 28 |
| 6 | Purchase | 38 |
| 7 | Purchased | 6 |
| 8 | Hospital HTA | 0 |
| 9 | Hospitals HTA | 0 |
| 10 | Hospitals Health Technology Assessment | 0 |
| 11 | Hospital Health Technology Assessment | 0 |
| 12 | Total | 103 |

**EconLit via ProQuest**

| Set# | Searched for | Results |
| --- | --- | --- |
| S1 | ti(Hospital OR Hospitals OR Hospice OR Hospices) OR ab(Hospital OR Hospitals OR Hospice OR Hospices) | 6700 |
| S2 | ti(Device OR Devices OR Equipment OR Supply OR Supplies) OR ab(Device OR Devices OR Equipment OR Supply OR Supplies) | 64074 |
| S3 | ti(Procure OR Procurement OR Procuring OR Procured OR Purchase OR Purchasing OR Purchased OR HTA OR HTAs OR miniHTA or miniHTAs OR (Technolog* N/1 Appais*) OR (Technolog* N/1 Assess*) OR (Technolog* N/1 Evaluat*)) OR ab(Procure OR Procurement OR Procuring OR Procured OR Purchase OR Purchasing OR Purchased OR HTA OR HTAs OR miniHTA or miniHTAs OR (Technolog* N/1 Appais*) OR (Technolog* N/1 Assess*) OR (Technolog* N/1 Evaluat*)) | 23950 |
| S4 | (ti(Hospital OR Hospitals OR Hospice OR Hospices) OR ab(Hospital OR Hospitals OR Hospice OR Hospices)) AND (ti(Device OR Devices OR Equipment OR Supply OR Supplies) OR ab(Device OR Devices OR Equipment OR Supply OR Supplies)) AND (ti(Procure OR Procurement OR Procuring OR Procured OR Purchase OR Purchasing OR Purchased OR HTA OR HTAs OR miniHTA OR miniHTAs OR (Technolog* NEAR/1 Appais*) OR (Technolog* NEAR/1 Assess*) OR (Technolog* NEAR/1 Evaluat*)) OR ab(Procure OR Procurement OR Procuring OR Procured OR Purchase OR Purchasing OR Purchased OR HTA OR HTAs OR miniHTA OR miniHTAs OR (Technolog* NEAR/1 Appais*) OR (Technolog* NEAR/1 Assess*) OR (Technolog* NEAR/1 Evaluat*))) | 40 |

**Embase via Ovid SP <1974 to 2020 Week 32>**

1	exp *Health Care Facility/ or exp *Hospital/ or *Hospice/ or *Hospital Department/ or exp *"Hospital Subdivisions and Components"/ or exp *Hospital Equipment/ or *Hospital Purchasing/ or (Hospital or Hospitals or Hospice*).ti,ab. (1993371)

2	exp *Medical Device/ or exp *Hospital Equipment/ or *Dental Technology/ or exp *Medical Technology/ or *Surgical Technology/ or (Device* or Equipment* or Supply or Supplies).ti,ab. (1662432)

3	*Hospital Purchasing/ or exp *Purchasing/ or *Biomedical Technology Assessment/ or (Procur* or Purchas* or HTA or HTAs or miniHTA or miniHTAs or (Technolog* adj1 (Appais* or Assess* or Evaluat*))).ti,ab. (83007)

4	1 and 2 and 3 (4837)

5	limit 4 to (conference abstracts or embase) (2582)

**Google Scholar**

allintitle: hospital|hospitals|hospice|hospices device|devices|equipment|supply|suplies|technology|technologies procurement|procure|procuring|procured|purchasing|purchase|purchased|HTA|"Technology Assessment"|minihta

340

**Google**

allintitle: hospital|hospitals|hospice|hospices device|devices|equipment|supply|suplies|technology|technologies procurement|procure|procuring|procured|purchasing|purchase|purchased|HTA|"Technology Assessment"|minihta

91

**HMIC Health Management Information Consortium via Ovid SP <1979 to July 2020>**

1	exp Hospitals/ or exp Hospital Departments/ or Hospices/ or exp Hospital Supplies/ or exp Hospital Equipment/ or (Hospital or Hospitals or Hospice*).ti,ab. (57617)

2	Equipment/ or Supplies/ or Health Service Equipment/ or Health Service Supplies/ or exp Hospital Supplies/ or exp Hospital Equipment/ or Medical Equipment/ or Medical Supplies/ or Ambulance Equipment/ or Ventilation Equipment/ or exp Surgical Equipment/ or exp Medical Instruments/ or Health Technology/ or exp Medical Technology/ or (Device* or Equipment* or Supply or Supplies).ti,ab. (14344)

3	Procurement/ or Purchasing/ or Baby Buying/ or Bulk Purchasing/ or Central Purchasing/ or Contract Purchasing/ or Joint Purchasing/ or Locality Purchasing/ or Total Purchasing/ or Purchasing Plans/ or Total Purchasing Projects/ or Purchasing Policies/ or exp Purchasing Officers/ or Purchasing Intelligence/ or Health Technology Assessment/ or (Procur* or Purchas* or HTA or HTAs or miniHTA or miniHTAs or (Technolog* adj1 (Appais* or Assess* or Evaluat*))).ti,ab. (9457)

4	1 and 2 and 3 (283)

**IEEE Xplore digital library**

| Hospital* | AND | Device* | AND | Procur* | 0 |
| --- | --- | --- | --- | --- | --- |
|  |  |  | AND | Purchas* | 0 |
|  |  |  | AND | HTA* | 0 |
|  |  |  | AND | miniHTA* | 0 |
|  |  |  | AND | "Technology Assessment" | 1 |
|  | AND | Equipment | AND | Procur* | 1 |
|  |  |  | AND | Purchas* | 0 |
|  |  |  | AND | HTA* | 0 |
|  |  |  | AND | miniHTA* | 0 |
|  |  |  | AND | "Technology Assessment" | 0 |
|  | AND | Supply | AND | Procur* | 0 |
|  |  |  | AND | Purchas* | 0 |
|  |  |  | AND | HTA* | 0 |
|  |  |  | AND | miniHTA* | 0 |
|  | AND | Supplies | AND | "Technology Assessment" | 0 |
|  |  |  | AND | Procur* | 0 |
|  |  |  | AND | Purchas* | 0 |
|  |  |  | AND | HTA* | 0 |
|  |  |  | AND | miniHTA* | 0 |
|  |  |  | AND | "Technology Assessment" | 0 |
|  | AND | Technolog* | AND | Procur* | 1 |
|  |  |  | AND | Purchas* | 0 |
|  |  |  | AND | HTA* | 0 |
|  |  |  | AND | miniHTA* | 0 |
|  |  |  | AND | "Technology Assessment" | 3 |

**INAHTA HTA database**

("Health Facilities"[mh] OR "Hospitals"[mhe] OR "Hospital Departments"[mhe] OR "Equipment and Supplies, Hospital"[mhe] OR "Purchasing, Hospital"[mhe] OR (Hospital* OR Hospice*)[Title] OR (Hospital* OR Hospice*)[abs]) AND ("Equipment and Supplies"[mh] OR "Equipment and Supplies, Hospital"[mhe] OR "Biomedical Technology"[mhe] OR (Device* OR Equipment* OR Supply OR Supplies)[Title] OR (Device* OR Equipment* OR Supply OR Supplies)[abs]) AND ("Purchasing, Hospital"[mhe] OR "Value-Based Purchasing"[mh] OR "Technology Assessment, Biomedical"[mhe] OR (Procur* OR Purchas* OR HTA* OR miniHTA* OR "Technology Assessment")[Title] OR (Procur* OR Purchas* OR HTA* OR miniHTA* OR "Technology Assessment")[abs])	43

**Ovid MEDLINE(R) and Epub Ahead of Print, In-Process & Other Non-Indexed Citations and Daily <1946 to August 12, 2020>**

1	*Health Facilities/ or exp *Hospitals/ or exp *Hospital Departments/ or exp *"Equipment and Supplies, Hospital"/ or exp *Purchasing, Hospital/ or (Hospital or Hospitals or Hospice*).ti,ab. (1281022)

2	*"Equipment and Supplies"/ or exp *"Equipment and Supplies, Hospital"/ or exp *Biomedical Technology/ or (Device* or Equipment* or Supply or Supplies).ti,ab. (674647)

3	exp *Purchasing, Hospital/ or *Value-Based Purchasing/ or exp *Technology Assessment, Biomedical/ or (Procur* or Purchas* or HTA or HTAs or miniHTA or miniHTAs or (Technolog* adj1 (Appais* or Assess* or Evaluat*))).ti,ab. (60766)

4	1 and 2 and 3 (2677)

**NHS EED and HTA via CRD**

Any Field	Device* OR Equipment* OR Supply OR Supplies AND

Any Field	Hospital* OR Hospice* AND

Any Field	Purchas* OR Procur* OR "Technology Assessment" OR HTA*

In NHS EED and HTA

381

**Open Access Theses and Dissertations**

title:(procurement OR procure OR procuring OR procured OR purchase OR purchasing OR purchased OR hta OR "health technology assessment") AND title:(hospital OR hospitals OR hospice OR hospices) AND title:(device OR devices OR equipment OR supply OR supplies)

5 results

**ProQuest Dissertations & Theses A&I**

| Set# | Searched for | Results |
| --- | --- | --- |
| S1 | ti(Hospital OR Hospitals OR Hospice OR Hospices) OR ab(Hospital OR Hospitals OR Hospice OR Hospices) | 50088 |
| S2 | ti(Device OR Devices OR Equipment OR Supply OR Supplies) OR ab(Device OR Devices OR Equipment OR Supply OR Supplies) | 247605 |
| S3 | ti(Procure OR Procurement OR Procuring OR Procured OR Purchase OR Purchasing OR Purchased OR HTA OR HTAs OR miniHTA or miniHTAs OR (Technolog* N/1 Appais*) OR (Technolog* N/1 Assess*) OR (Technolog* N/1 Evaluat*)) OR ab(Procure OR Procurement OR Procuring OR Procured OR Purchase OR Purchasing OR Purchased OR HTA OR HTAs OR miniHTA or miniHTAs OR (Technolog* N/1 Appais*) OR (Technolog* N/1 Assess*) OR (Technolog* N/1 Evaluat*)) | 32069 |
| S4 | (ti(Hospital OR Hospitals OR Hospice OR Hospices) OR ab(Hospital OR Hospitals OR Hospice OR Hospices)) AND (ti(Device OR Devices OR Equipment OR Supply OR Supplies) OR ab(Device OR Devices OR Equipment OR Supply OR Supplies)) AND (ti(Procure OR Procurement OR Procuring OR Procured OR Purchase OR Purchasing OR Purchased OR HTA OR HTAs OR miniHTA OR miniHTAs OR (Technolog* NEAR/1 Appais*) OR (Technolog* NEAR/1 Assess*) OR (Technolog* NEAR/1 Evaluat*)) OR ab(Procure OR Procurement OR Procuring OR Procured OR Purchase OR Purchasing OR Purchased OR HTA OR HTAs OR miniHTA OR miniHTAs OR (Technolog* NEAR/1 Appais*) OR (Technolog* NEAR/1 Assess*) OR (Technolog* NEAR/1 Evaluat*))) | 153 |

**Scopus**

#4	#1 AND #2 AND #3	2,014

#3	( TITLE ( procur* OR purchas* OR hta OR htas OR minihta OR minihtas OR ( technolog* PRE/1 appais* ) OR ( technolog* PRE/1 assess* ) OR ( technolog* PRE/1 evaluat* ) ) OR ABS ( procur* OR purchas* OR hta OR htas OR minihta OR minihtas OR ( technolog* PRE/1 appais* ) OR ( technolog* PRE/1 assess* ) OR ( technolog* PRE/1 evaluat* ) ) )	231,105

#2	( TITLE ( device* OR equipment* OR supply OR supplies ) OR ABS ( device* OR equipment* OR supply OR supplies ) )	3,225,577

#1	( TITLE ( hospital OR hospitals OR hospice OR hospices ) OR ABS ( hospital OR hospitals OR hospice OR hospices ) )	1,449,788

**Web of Science databases**

- Science Citation Index Expanded (SCI-EXPANDED) --1900-present
- Conference Proceedings Citation Index- Science (CPCI-S) --1990-present
- Emerging Sources Citation Index (ESCI) --2015-present

(TI=(Hospital OR Hospitals OR Hospice OR Hospices) OR AB=(Hospital OR Hospitals OR Hospice OR Hospices)) AND (TI=(Device* OR Equipment* OR Supply OR Supplies) OR AB=(Device* OR Equipment* OR Supply OR Supplies)) AND (TI=(Procur* OR Purchas* OR HTA OR HTAs OR miniHTA OR miniHTAs OR (Technolog* NEAR/1 Appais*) OR (Technolog* NEAR/1 Assess*) OR (Technolog* NEAR/1 Evaluat*) ) OR AB=(Procur* OR Purchas* OR HTA OR HTAs OR miniHTA OR miniHTAs OR (Technolog* NEAR/1 Appais*) OR (Technolog* NEAR/1 Assess*) OR (Technolog* NEAR/1 Evaluat*) ))

Indexes=SCI-EXPANDED, CPCI-S, ESCI Timespan=All years	804

**Zetoc Conference Search**

| **Search** | **Hits** | **Search terms** |
| --- | --- | --- |
| 1 | 1 | tip:Procure Hospital |
| 2 | 0 | tip:Procured Hospital |
| 3 | 5 | tip:Procurement Hospital |
| 4 | 0 | tip:Procuring Hospital |
| 5 | 0 | tip:Procure Hospitals |
| 6 | 0 | tip:Procured Hospitals |
| 7 | 2 | tip:Procurement Hospitals |
| 8 | 0 | tip:Procuring Hospitals |
| 9 | 1 | tip:Purchase Hospital |
| 10 | 0 | tip:Purchased Hospital |
| 11 | 3 | tip:Purchasing Hospital |
| 12 | 0 | tip:Purchase Hospitals |
| 13 | 0 | tip:Purchased Hospitals |
| 14 | 3 | tip:Purchasing Hospitals |
| 15 | 2 | tip:Health Technology Assessment Hospital |
| 16 | 3 | tip:HTA Hospital |
| 17 | 0 | tip:Health Technology Assessment Hospitals |
| 18 | 0 | tip:HTA Hospitals |
| Total | 20 |  |
